# Hippocampal subfield volume in relation to cerebrospinal fluid Amyloid-ß in early Alzheimer’s disease: Diagnostic Utility of 7T MRI

**DOI:** 10.1101/2024.10.24.24315913

**Authors:** Oluwatobi F Adeyemi, Penny Gowland, Richard Bowtell, Olivier Mougin, Akram A. Hosseini

## Abstract

**INTRODUCTION:** Alzheimer’s disease (AD) is a neurodegenerative condition characterised by amyloid plaque accumulation and neurofibrillary tangles. Early detection is essential for effective intervention, but current diagnostic methods that enable early diagnosis in clinical practice rely on invasive or costly biomarker scanning. This study aimed to explore the utility of 7T MRI in assessing hippocampal subfield volumes and their correlation with cerebrospinal fluid (CSF) biomarkers in prodromal AD. METHODS: Fifty-six participants, including AD patients and healthy controls, underwent 7T MRI scanning. Automated segmentation delineated hippocampal subfield volumes, with subsequent normalization to whole brain volume.

**RESULTS:** Significant differences in hippocampal and subfield volumes were observed in prodromal AD patients, even when they did not exhibit high MTA scores on 3T MRI or show any whole brain volume loss. Additionally, the volume of the entorhinal cortex (ERC) correlated significantly with CSF amyloid-β levels, suggesting ERC’s potential as a proxy CSF amyloid-ß measurement. Conversely, no significant associations were found between CSF 181-phospho-tau or total tau levels and any hippocampal subfield volumes.

**DISCUSSION:** These findings show the potential use of 7T MRI, particularly in ERC assessment, as a biomarker for early AD identification. Further validation studies are warranted to confirm these results and elucidate the relationship of ERC volume with CSF biomarkers.

## BACKGROUND

Alzheimer’s disease (AD) is the leading cause of dementia. It is a multifactorial condition influenced by various genetic, environmental, and molecular factors ^1,2^. AD is a progressive neurodegenerative disease that is accompanied by the presence of neurofibrillary tangles and the accumulation of amyloid plaques ^3^, for which the main constituent is the amyloid-ß (Aß) protein. Aß is first deposited in the neocortex, and then spreads into the hippocampus, amygdala, and cingulate gyrus ^4^. However, neurofibrillary tangles occur in trans-entorhinal and entorhinal cortex before invading the subiculum, Cornus Ammonis CA1, and then CA2 and CA3 hippocampal subfields, and finally neocortex ^5,6^. Post-mortem examination of brain tissue is the gold standard for the diagnosis of AD, and clinical diagnosis based on cognitive symptoms provides suboptimal sensitivity and specificity^7^.

Early identification of AD is key for consideration of Disease-Modifying-Treatments that are in the final stages of investigation ^8–10^. Pathological changes in Aß and tau can occur more than a decade before the onset of dementia ^11,12^. Aß measurements in the cerebrospinal fluid (CSF) are known to correlate positively with cognitive scores whereas Phosphorylated-Tau and total Tau levels in the CSF correlate negatively with Mini-Mental State Examination (MMSE) scores ^13^. The presence of Aß with or without tau proteins, combined with radiological topography of neurodegeneration, currently provides the best accuracy for the diagnosis of AD in patients who present with subjective cognitive impairment ^14–17^. Aß, tau- pathology and neurodegeneration (ATN categories) are diagnostic markers to identify AD at the early stages ^18,19^. Aß pathology can be detected using an Amyloid PET scan, or by obtaining CSF through a diagnostic lumbar puncture to test for Aß and tau levels. While both methods are highly accurate for detecting AD, they are invasive, or expensive, or both. Furthermore, diagnostic CSF biomarker assessments does not provide the topographic information about specific brain regions affected.

Medial Temporal Atrophy (MTA) scores, which characterise the atrophy of the whole hippocampus on structural MRI, form an established neuroimaging biomarker for AD in clinical practice ^20^. However the atrophy of the whole hippocampus, that can be identified using clinical MRI at 1.5 or 3T, is a late marker of neurodegeneration in AD when the disease is advanced ^21^.

Ultrahigh field 7T MRI offers a non-invasive technique that is free of ionizing radiation which can produce images of high contrast and high spatial resolution. Further, MRI at 7T can provide a direct measure of local neurodegeneration ^22,23^, and validated pathological confirmation of hippocampal volume ^24^, whereas the CSF pathological state of Aβ and tau do not provide information on focal neuronal loss. Structural MRI at 7T has been shown to be reproducible across multiple sites ^25^ and can be used to measure the volume of hippocampal subfields ^26,27^.

We have previously demonstrated that 7T MRI has the ability to measure differences in hippocampal subfield volume compared to healthy controls in patients with early AD who do not exhibit diagnostic high MTA scores on 3T MRI ^28^. Further, we observed a reduction in hippocampal subfield volume in these patients with early AD, even without a loss of total brain volume, which correlated with their clinical amnestic impairment ^28^. Here, we aim to utilise 7T MRI to precisely measure the volume of each hippocampal subfield in the early stages of AD and to correlate these measurements with Aβ and tau levels in CSF, as well as with levels of cognitive decline. Our study investigates whether 7T MRI can enhance diagnostic accuracy for AD in its early stages, as defined by the ATN criteria, before hippocampal atrophy is evident on clinical MRI.

We hypothesise that lower subfield volume will correspond to more abnormal levels of amyloid/tau status in the CSF and/or worse performance on the cognitive assessments.

## METHODS

### Participants

Fifty-six participants (31 AD patients and 25 healthy participants), aged between 40 and 79 years, were enrolled through prospective recruitment. The participants with AD fulfilled the A+T+ or A+T- criteria on ATN framework for the AD diagnosis, confirmed by the presence of Aß, with or without tau, in the CSF of symptomatic patients who had mild cognitive impairment ^29^. Participants with clear evidence of hippocampal atrophy (i.e. MTA scale > 2) on their clinical MRI at 3T were considered to have advanced neurodegeneration and were excluded from the 7T MRI study. The CSF Aß/tau measurement cut off values were based on the reference values (Aß_42_ range 627 – 1322 pg/ml; total tau range 146 -595 pg/ml; and Thr181-phosphorylated-tau range below 68 pg/ml)^30^ as previously reported ^31^. The time between CSF sampling and 7T MRI was less than 3 months. Three individuals were also scanned where the interval ranged from 23 to 51 months, but they were excluded from further analysis because of the significant separation in time of the two different measurements.

Inclusion criteria for the healthy control group included cognitive performance within 1.5 SD of normal in all cognitive tests ^32^. As described previously, participants were assessed cognitively using the Uniform Data set (UDSNB3.0) which includes the MoCA (Montreal Cognitive Assessment ) test ^28^. The exclusion criteria included contraindications to Ultrahigh Field MRI, dementia (defined as loss of at least one functional ability according to the DSM- 5 Criteria ^29,33^) and incompetency to consent.

This project received favourable opinion from the East Midlands-Nottingham 2 research Ethics committee (reference: 20/EM/0023) and the UK Health Research Authority (IRAS:276174). The committee is constituted in accordance with the Governance Arrangement for research Ethics committee and complies fully with the Standard Operating Procedures for Research Ethics Committee in the UK.^31^ (www.ClinicalTrials.gov ID NCT04992975).

## IMAGING PROTOCOL

The participants were scanned on a Philips Achieva 7T system using a Nova Medical (Wilmington MA, USA) single-channel transmit, 32-channel receive (1Tx32Rx) head coil at the Sir Peter Mansfield Imaging Centre at the University of Nottingham.

A whole-head, T_1_-weighted PSIR data set was acquired first with inversion times of 725/2150 ms; TE= 3.1 ms; TR= 6.9 ms; Field of view (FOV) 192 × 183 × 157 mm^3^; voxel size 0.55 × 0.55 ×0.55 mm^3^; receiver bandwidth of 300 Hz; total scan time 12:25 minutes.

T_2_-weighted images spanning the hippocampus were acquired using a TSE sequence: TE = 117 ms; TR = 5900ms; FOV 224 × 224 × 64 mm^3^; voxel size 0.38 × 0.39 ×1.5 mm^3^; receiver bandwidth 155 Hz; total scan time 04:43 minutes. The slice orientation was chosen so that the slices ran orthogonal to the longest axis of the hippocampus.

## IMAGE PROCESSING AND ANALYSIS

Automatic Segmentation of Hippocampus Subfield (ASHS) was used to segment the hippocampus from the PSIR and T_2_-weighted images ^34^ using an atlas created by the UMC Utrecht group (ashs_atlas_umcutrecht_7t_20170810) and provided by the NeuroImaging Tools and Resources Collaboratory (NITRC))^27^. In this atlas, as shown in Figure 1, the hippocampus is segmented into eight subfield ROIs: the Cornu Ammonis (CA) areas: CA1, CA2 and CA3, the hippocampal tail (TAIL), the Dentate Gyrus (DG), the Subiculum (SUB), the cyst and the Entorhinal Cortex (ERC). The cyst volume was omitted because this volume was very small or not detected in all participants.

**Figure 1:**
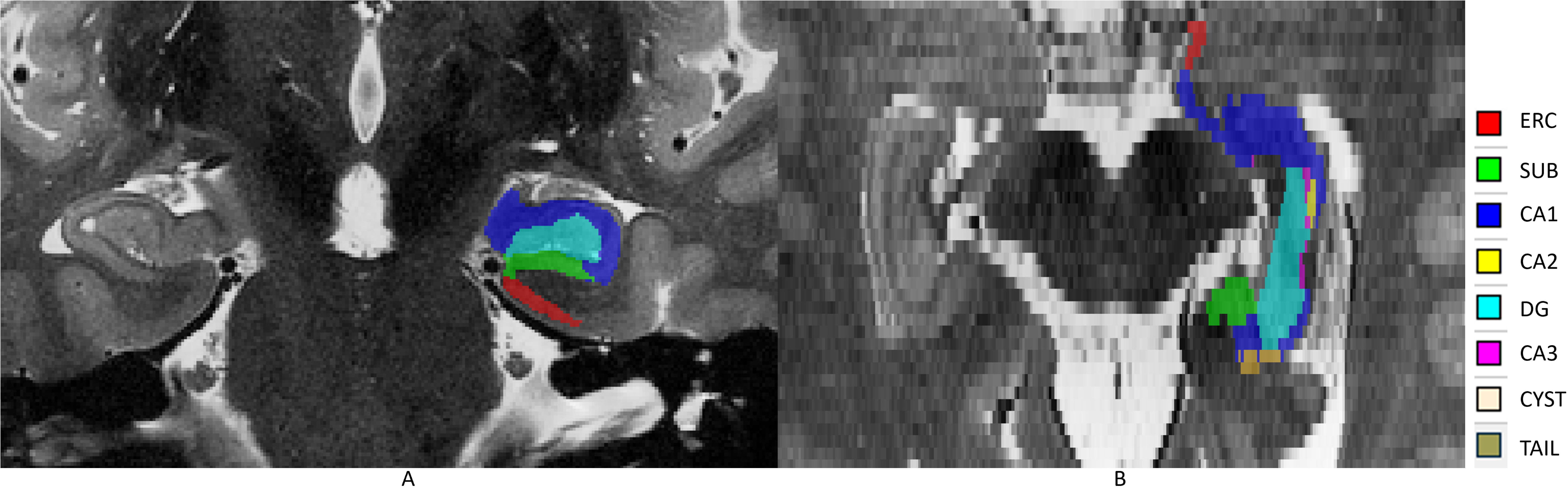
Segmentation of the hippocampus overlaid on the T2-weighted image for (A) Coronal view and (B) Axial view in one representative subject.

## PERCENTAGE VOLUME OF THE HIPPOCAMPAL SUBFIELDS

The volume of the whole brain differs between participants (with significant differences between male and female individuals), but we are interested in specific atrophy of the hippocampus. Therefore, we normalised the volume of the hippocampus to the volume of the whole brain. The whole brain volume (including the CSF and cerebellum) was extracted from the PSIR image data.

## STATISTICAL ANALYSIS

Statistical analysis was carried out using T-test and regression analysis for comparison between AD and age-matched healthy participants. The volumes of the left and right hippocampal subfields were combined to provide increased sensitivity. We conducted a statistical analysis to compare the volumes of the right and left hippocampus across the different subfields and the whole hippocampus in both the HC and AD groups. Our results revealed no significant difference between the two hemispheres for the entorhinal cortex (ERC), Cornu Ammonis regions 1-3 (CA1-3), dentate gyrus (DG), and the whole hippocampus, with p-values ranging from 0.194 to 0.99. However, a significant difference was observed in the hippocampal tail, with a p-value of 0.001.

The statistical differences in subfield volumes (CA1, CA2, CA3, ERC, TAIL, DG, and SUB) between the AD and HC groups were evaluated using SPSS (version 27) and adjusted for multiple comparisons using the Bonferroni correction across the seven hippocampal subfields analysed in this study. The analysis of the correlation between CSF measures and sub-field volumes was also subjected to Bonferroni correction to account for multiple comparisons.

## RESULTS

In addition to the three participants excluded due to the extended time between CSF measurement and MRI, data from three further participants with AD were excluded: one due to inability to undergo imaging due to claustrophobia, one due to image degradation related to motion artefacts and one participant was excluded because clinical images revealed an MTA scale of 3. Hence, the analysis was performed on data from 25 patients with AD and 25 age-matched control participants. The participant demographics are shown in Table 1.

**Table 1:** Demographics of the participants with Alzheimer’s disease (AD) and healthy Controls (HC).

|  | **AD Group** | **HC Group** | **P-value** |
| --- | --- | --- | --- |
| Number | 25 | 25 |  |
| Age (Mean, Range) | 60 (42-76) | 62 (52-79) | 0.436 |
| Sex (Male, Female) | 33 %, 67% | 44%, 56% | 0.643 |
| Mean MoCA score (Mean, Range) | 12 (4-22) | 27(23-29) | <0.001 |
| Smoker | 5(19%) | 9(39%) | 0.532 |
| Alcohol (only mild to moderate consumption) | 12(44%) | 12(52%) | 0.41 |
| Time between CSF sampling and MRI (AD) | 1-51 months |  |  |

There was no significant difference in risk factors such as smoking and alcohol habits between the AD and control participants.

Visual examination was employed to assess the accuracy of the automated hippocampal segmentation. In instances where segmentation quality was compromised by motion artifacts during scanning, participants were offered the opportunity for a repeat scan (N=3). However, one participant’s image data showed excessive motion artifacts in both the initial and repeat scans, resulting in their exclusion from the study, as previously noted.

Figure 2A shows that total brain volume was smaller in the AD group as compared to the healthy controls, although this difference was not significant. Figure 2B shows that there was a significant reduction in the volume of the whole hippocampus in patients with AD (p < 0.001).

**Figure 2:**
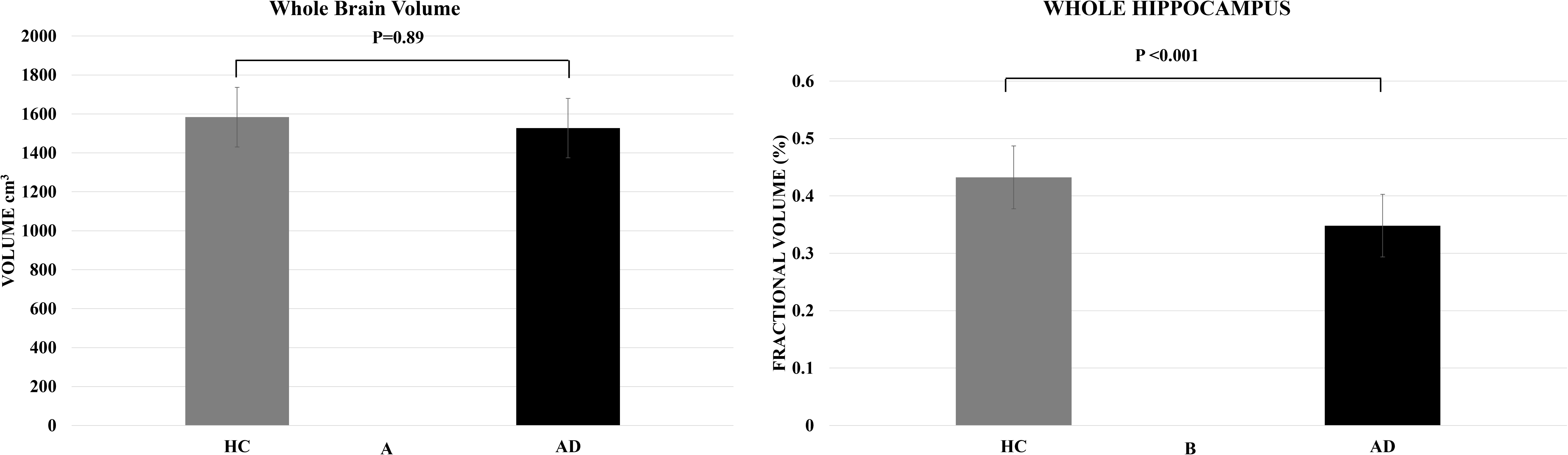
(A)The absolute volume of the whole brain averaged over the Alzheimer’s disease and healthy control participant groups. There was no statistical significant between the groups. (B) The fractional volume of the whole hippocampus averaged over the Alzheimer’s disease and healthy control participant groups. There was a statistically significant reduction in hippocampal volume in the AD patients compared to the controls. Error bars indicate inter-subject standard deviation.

Figure 3 shows that there was a significant reduction in the fractional volume of the ERC (p < 0.001), DG (p < 0.001), CA1 (p=0.016), CA2 (p < 0.001) and Tail (p=0.003) but SUB and CA3 subfield did not attain statistical significance in AD patients compared to controls with p = 0.516 and 0.235 respectively. These findings are consistent with our previous observations reported in ^28^. However, in this study, we have included a larger sample size, which including 7 additional HC and 1 more AD participant. Detailed comparisons and the expanded dataset can be found in Appendix Table A2.

**Figure 3:**
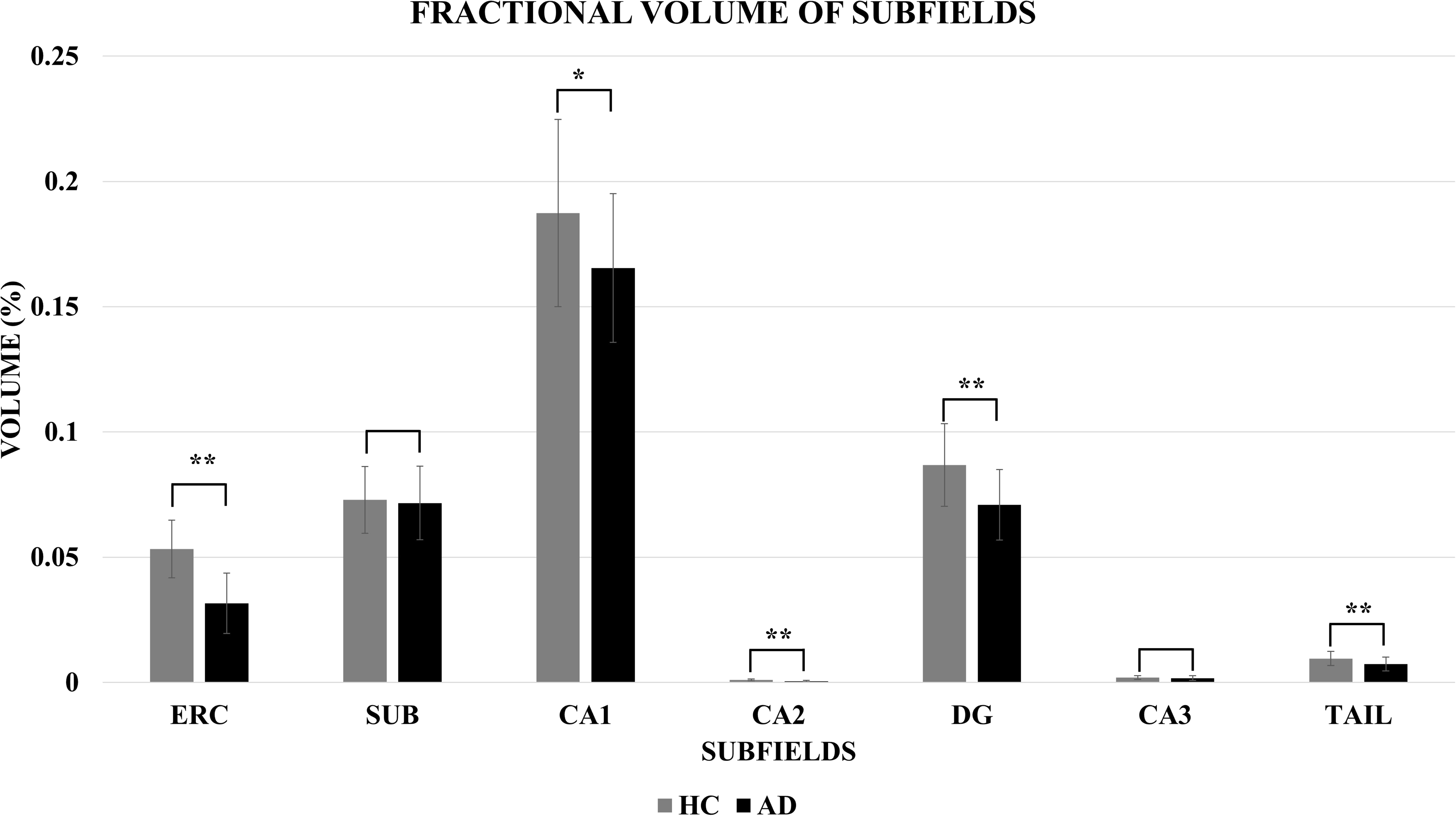
The volume of the hippocampal subfields as a percentage of whole brain volume averaged over the Alzheimer’s disease and healthy control participants. Error bars indicate intersubject standard deviation; *indicates significant p-value and ** indicate significance with Bonferroni correction.

Figure 4 and Table 2 show the relationship between the fractional volumes of the hippocampus subfields and CSF Aβ_42_ (which was only measured for AD subjects). All subfields showed a positive correlation between fractional volume and levels of Aβ_42_ in the CSF (i.e. smaller fractional volume for lower levels of Aβ_42_), but this only reached significance for the ERC (R= 0.596; p=0.002). This relation was still significant after Bonferroni correction was carried out. Lower ERC volume was also significantly associated with impaired cued memory in the MoCA test. Details of the cognitive assessments in relation to volume of hippocampal subfields are shown in the Appendix table A1.

**Figure 4:**
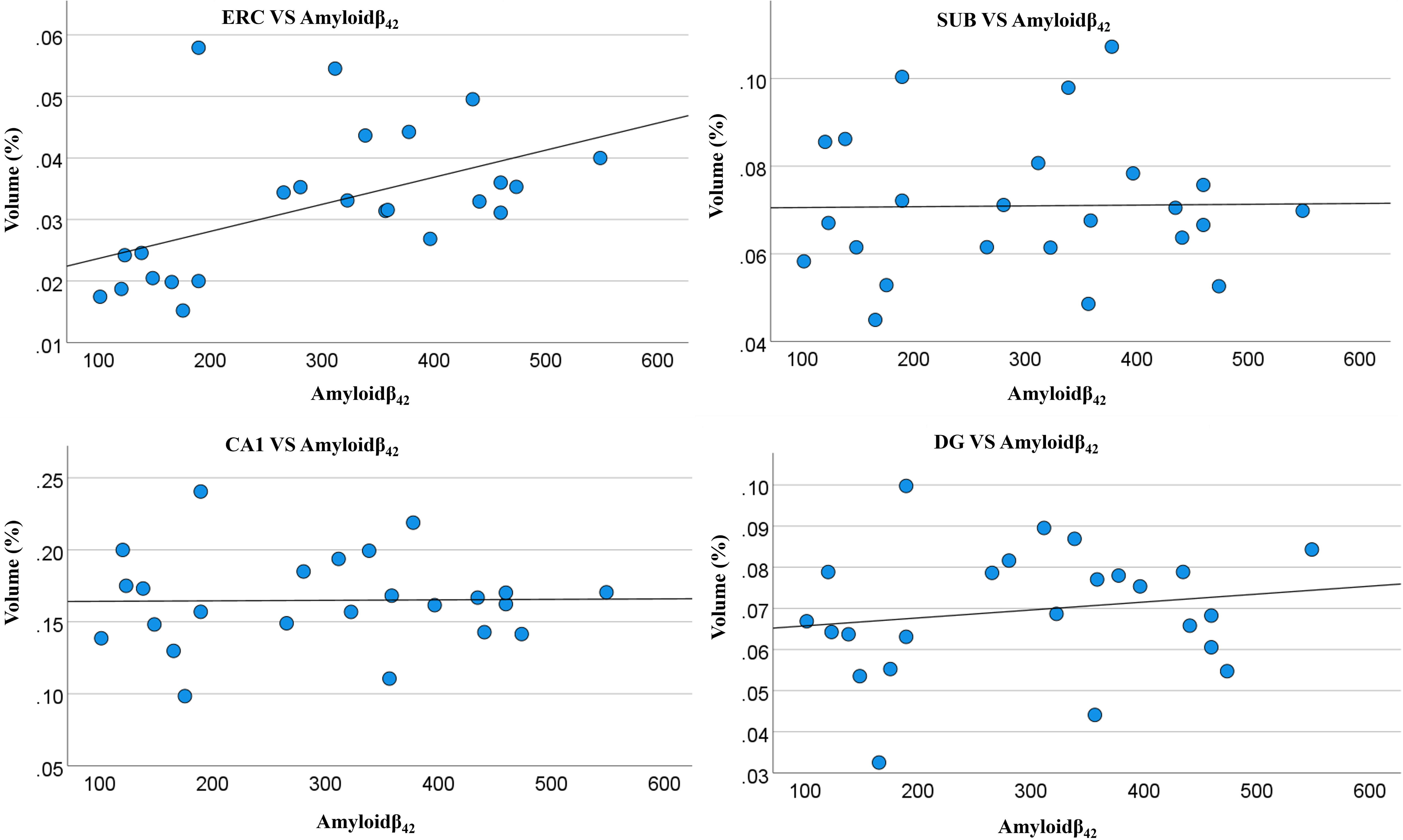
Relationship between the fractional volume of some of the hippocampal subfields for AD participants and the value of CSF amyloid-β42, with linear trend lines included.

**Table 2:** shows the Spearman’s rho correlation analysis for associations between hippocampal subfield volumes and abnormal CSF proteins in AD participants. *Indicates significant p-value and ** indicate significance with Bonferroni correction.

| Correlations |  |  |  |  |
| --- | --- | --- | --- | --- |
| Subfields | | Amyloid-$\beta_{42}$ | 181-Phosphorylated-tau | total tau |
| ERC | Correlation Coefficient | .596** | 0.029 | -0.144 |
|  | P-Value | 0.002 | 0.902 | 0.533 |
| SUB | Correlation Coefficient | 0.029 | 0.110 | -0.070 |
|  | P-Value | 0.894 | 0.634 | 0.763 |
| CA1 | Correlation Coefficient | -0.001 | -0.034 | -0.258 |
|  | P-Value | 0.995 | 0.884 | 0.258 |
| CA2 | Correlation Coefficient | 0.014 | .518* | 0.358 |
|  | P-Value | 0.950 | 0.023 | 0.132 |
| DG | Correlation Coefficient | 0.136 | 0.113 | -0.052 |
|  | P-Value | 0.526 | 0.626 | 0.823 |
| CA3 | Correlation Coefficient | 0.137 | 0.039 | -0.169 |
|  | P-Value | 0.523 | 0.867 | 0.464 |
| TAIL | Correlation Coefficient | -0.055 | -0.131 | -0.312 |
|  | P-Value | 0.799 | 0.571 | 0.169 |

ERC: entorhinal cortex; SUB: Subiculum; CA1-3: Cornu Ammonis regions 1-3, DG: dentate gyrus.

After Bonferroni correction, no significant associations were observed between the fractional volumes of the hippocampal subfields and the levels of CSF total tau or CSF 181- Phosphorylated tau, as detailed in Table 2. An initially significant correlation found for Phospho-tau in CA2, was no longer significant following the multiple correction analysis.

## DISCUSSION

In this study, we utilised ultrahigh field, 7T PSIR and T_2_-Weighted images to measure the volume of the hippocampus and its subfields in patients diagnosed with biological amyloid-status Alzheimer’s disease. These patients were categorised according to the ATN classifications as A+T-N- or A+T+N- ^29^, but they did not meet the AD diagnostic criteria for medial temporal atrophy (i.e., MTA scale 2 or above) on their clinical MRI data acquired at 3T. We compared the measured volumes to the levels of Aβ_42_ and tau in the CSF in patients with AD during the prodromal phase, when standard clinical MRI was not diagnostic.

In prodromal AD, we observed a significant decrease in the fractional volume of the whole hippocampus, as well as fractional reductions in the volumes of the ERC, DG, CA1, CA2 and Tail subfields (p<0.001) compared to age-matched controls ^28^.

Furthermore, in participants with prodromal AD, the volume of the ERC subfield (as a fraction of total brain volume) was positively correlated with the measured value of Aβ_42_ in the CSF (R^2^= 0.596 and p = 0.002), and this correlation remained significant after Bonferroni correction. This correlation reflects the linear relationship between abnormal CSF Aβ_42_ and ERC volume loss. These findings suggest that 7T MRI might serve as a proxy measure of CSF Aβ_42_, potentially offering an additional or alternative non-invasive biomarker with diagnostic and monitoring value for AD.

Previous neuroimaging studies using standard MRI at 3T and 1.5 T have consistently reported absolute hippocampal volume loss in AD ^34–36^. In this study, we examined patients with prodromal AD who did not have significant reduction in total brain volume, compared to age-matched healthy controls (p= 0.89; Figure 2B). Despite this, using 7T MRI, we were able to identify hippocampal volume loss in these patients. Our findings suggests that 7T MRI may outperform standard MRI (at 3 or 1.5T) as a potential marker for early identification of atrophy, thus aiding the diagnosis of AD.

It has been previously reported that the level of CSF Aβ_42_ declines during the preclinical stage of AD ^37^, and correlates with delays in memory tasks in patients with Mild Cognitive Impairment (MCI) ^38^. Our prior research has shown a significant association between the loss of hippocampal subfields and impaired memory tasks in patients with amyloid-positive prodromal AD ^28^. Further, we have found significant positive associations between the ERC volume and the level of CSF Aβ_42_, consistent with the notion that CSF Aβ_42_decline may serve as a marker of volume loss. The interaction between Aβ_42_ and 181-Phosphorylated-tau on hippocampal atrophy ^39^ and ERC atrophy ^40^ has previously been reported, and further longitudinal investigation indicated that Aβ status was linked to accelerated ERC atrophy in non-demented patients with positive 181-Phosphorylated-tau ^40^. In our study, six participants were classified as A+T- (i.e. with evidence of abnormal level of Aβ_42_, but normal level of total tau, and 181-Phosphorylated-tau in the CSF), whereas the remainder of participants were classified as A+T+ (i.e. abnormal Aβ_42_ and tau in the CSF). We investigated the correlation between the volume of the ERC and Aβ_42_ levels in the participants with A+T- and found no significant correlation in these 6 AD participants. While this finding could be related to a small sample size, there is evidence in the literature supporting that patients with abnormal Aβ but normal tau levels in the CSF might be a distinct subgroup within AD ^41,42^.

The ERC is known to play crucial roles in memory, navigation, and perception of time ^43,44^. Notably, it is also the region of the brain where the early histopathological changes of AD occurs before migrating along the hippocampus ^45^. The specificity of this correlation in the ERC suggests that this region may be particularly sensitive to amyloid-related damage. An early identification of ERC volume loss in the appropriate clinical context holds potential to be used as a surrogate measure of CSF Aβ_42_ along with the other biomarkers.

The measurement of tau proteins, specifically total tau and phosphorylated-tau, in CSF has been extensively studied as a potential biomarker for AD ^46–48^. Elevated levels of these proteins are often associated with neurodegeneration ^48^. Researchers have hypothesised that increased tau levels in CSF may correspond to greater neuronal damage, potentially leading to hippocampal atrophy ^49,50^.

The finding from our study that there was no significant association between CSF total tau, or CSF 181-Phosphorealted-tau, and the volume of hippocampal subfields implies a more complex relationship between CSF tau and hippocampal atrophy than previously thought. It is possible that the relationship between CSF tau and hippocampal atrophy evolves over time; tau pathology may initially lead to increased CSF tau levels, but this relationship may weaken as the disease progresses ^50,50–52^. Furthermore AD is a heterogeneous disease ^53^, and different neurodegenerative disorders such as “tauopathies” may exhibit variations in the type and distribution of tau pathology ^54^.

To use our findings for clinical translation, a larger sample size should be tested. This study was not aimed to assess the causality of hippocampal subfield atrophy in AD and further molecular and/or pathological studies are required to explore if there could be a direct relationship between the CSF pathological state of AD and hippocampal subfield atrophy. Although 7T MRI is less clinically available and more expensive than some other diagnostic modalities, the lack of radiation, non-invasiveness and enhanced sensitivity for a direct measurement of early structural changes in the brain suggests a potential clinical application of this modality which could be valuable as a proxy for amyloid-positive AD during the early stages of the disease, when the use of Disease-Modifying-Treatments need to be assessed.

By utilising 7T MRI to detect Aβ, clinicians can effectively address two pillars of the ATN scores (’A’ for amyloid and ’N’ for neurodegeneration) during a single imaging session. This comprehensive assessment aids in better understanding the disease progression, assisting in early diagnosis, and potentially enabling more targeted treatment strategies aimed at intervening in the disease process before significant neurodegeneration occurs.

In conclusion, the association between the volume of ERC and CSF Aβ_42_ suggests the potential for using high field-high resolution MRI as a biomarker for an early identification of AD. Larger studies are required to further evaluate the relationship between the ERC and Aβ_42_.

## Data Availability

Data is Available on request

## ACKNOWLEDGEMENTS

Chris Newby for statistics; Haley Morris, Ishani Hari, Hannah Sargisson, Clinical research Nurses at the Academic Neurology, NUH for data collection and recruitment, George Hutchinson for contribution to obtaining some images.

## CREDIT AUTHOR CONTRIBUTION STATEMENT

**Oluwatobi Folorunsho Adeyemi, Ph.D**: (Data curation; Formal analysis; Investigation; Methodology; Project administration; Software; Validation; Visualization; Writing – original draft; Writing – review & editing).

**Olivier Mougin, PhD**: (Formal analysis; Supervision; Validation; Visualization).

**Richard Bowtell, PhD**: (Methodology; Project administration; Resources; Supervision; Validation; Visualization; Writing – review & editing).

**Penny Gowland, PhD**: (Methodology; Project administration; Resources; Supervision; Validation; Visualization; Writing – review & editing).

**Akram A. Hosseini, MD, PhD, MRCP (UK)**: (Conceptualization; Funding acquisition; Methodology; Project administration; Resources; Supervision; Validation; Visualization; Writing – original draft; Writing – review & editing).

## CONSENT STATEMENT

All participants involved in this study provided informed consent. Participants were informed about the nature, purpose, procedures, potential risks, and benefits of the study. They were given the opportunity to ask questions and were assured that their participation was voluntary, with the option to withdraw at any time without any negative consequences. Written consent forms were obtained from all participants prior to their involvement in the study.

## APPENDIX

**Table A1:**
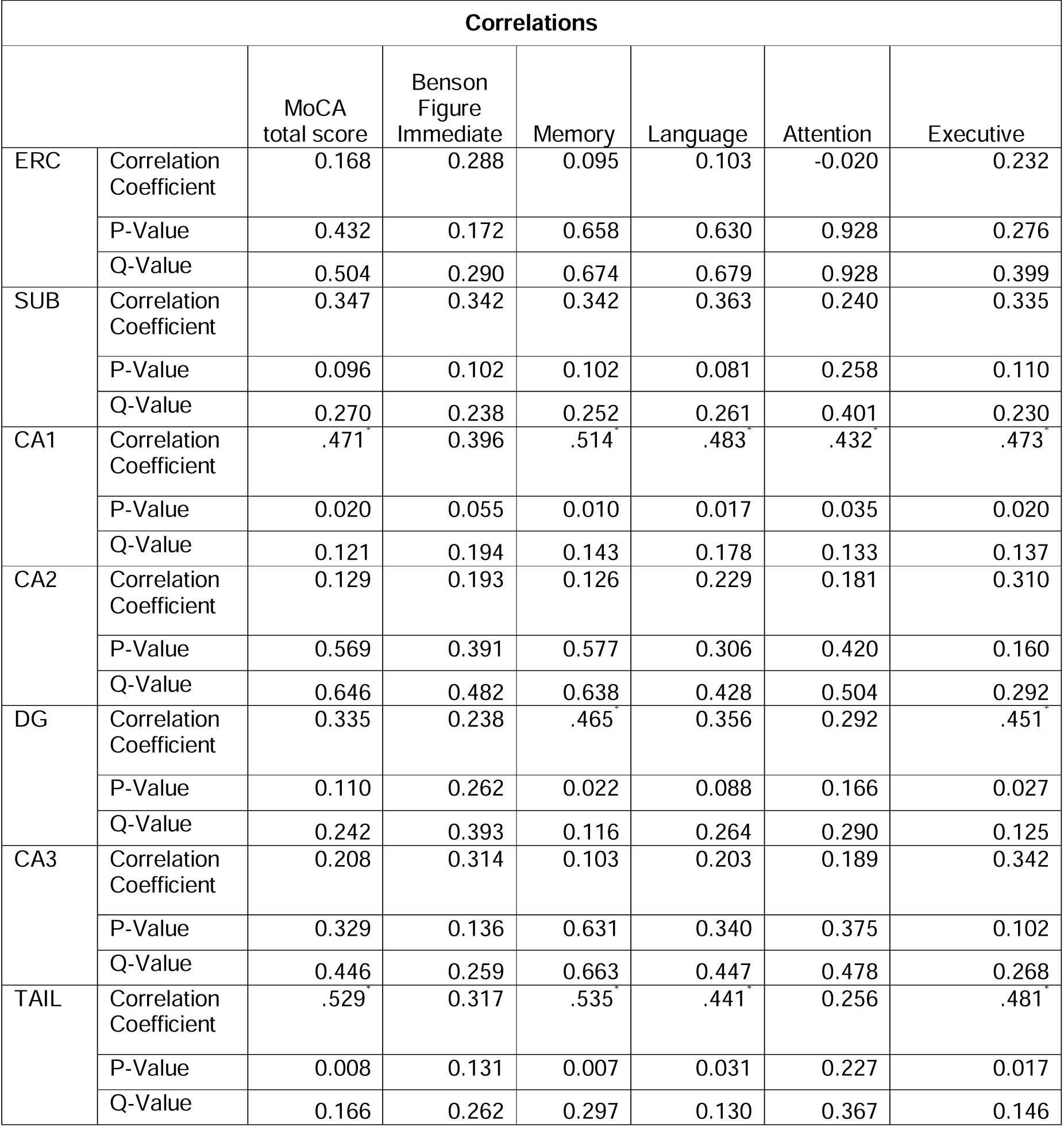
Spearman’s rho correlation coefficients for associations between hippocampal subfield volumes and cognitive scores in AD participants, the P-values, and the Q-values (P-values adjusted by Benjamini–Hochberg correction)

**Table A2:**
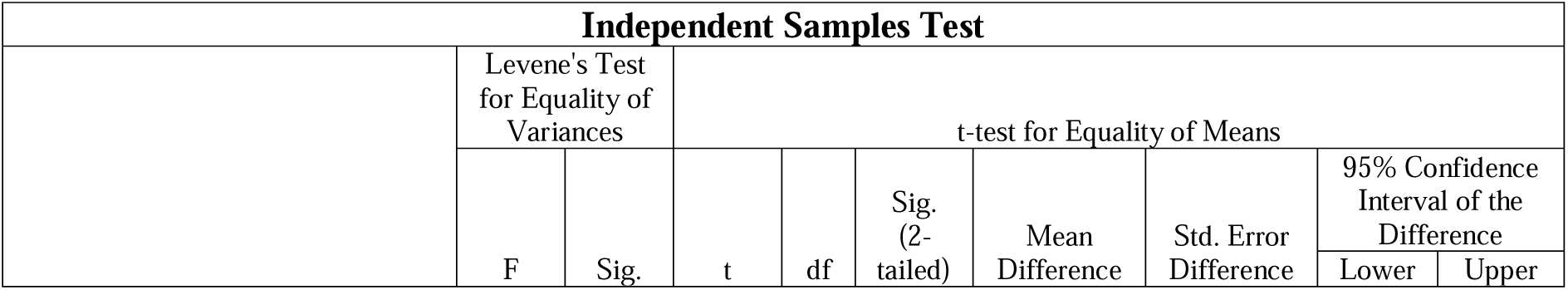
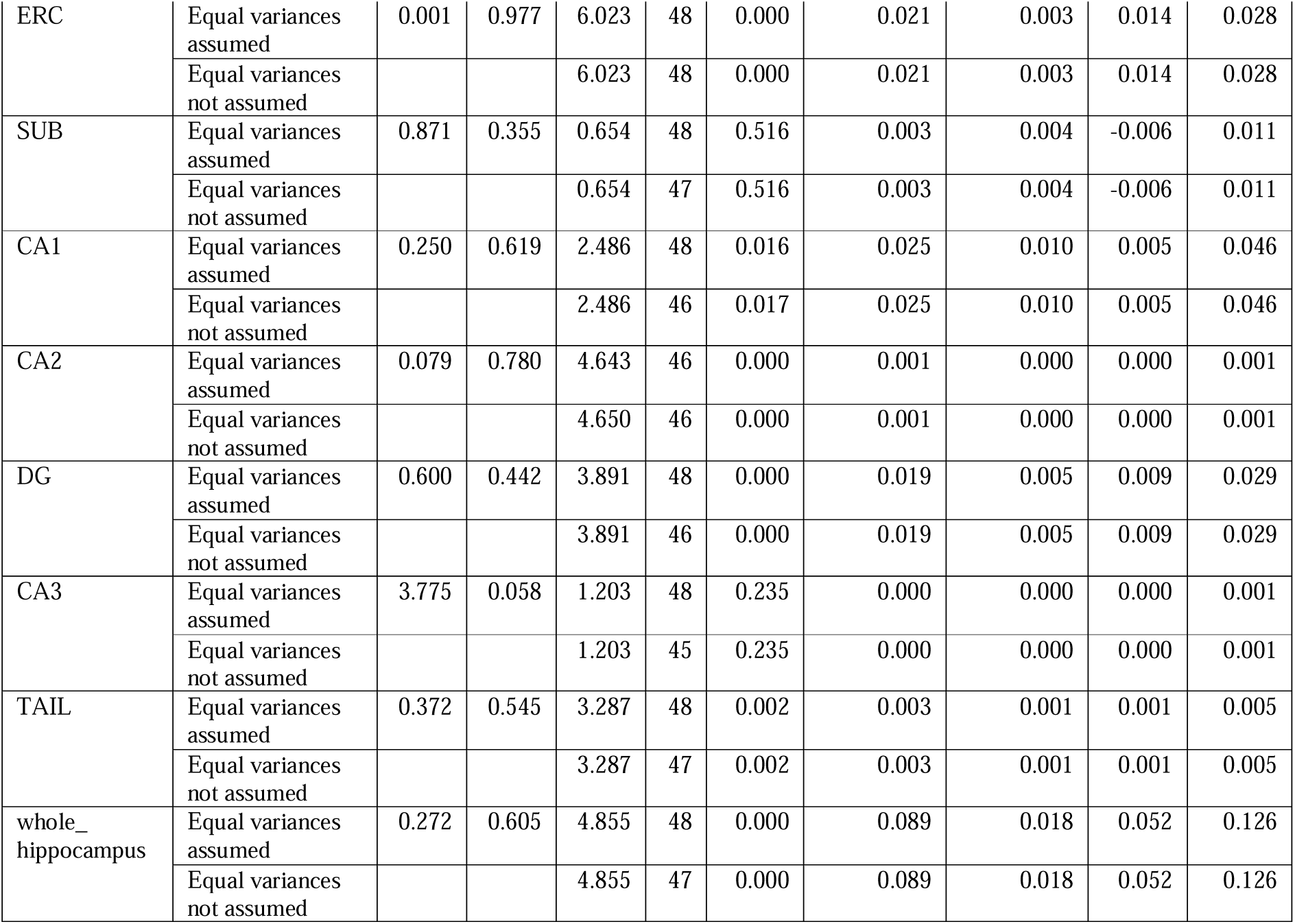
The results of the t-test comparing the hippocampal subfield volumes between Alzheimer’s disease (AD) and healthy control (HC) participants. A Bonferroni correction was applied to account for multiple comparisons, setting the adjusted p-value threshold at 0.006.

## REFERENCE

1. Iqbal K, Grundke-Iqbal I. Alzheimer’s disease, a multifactorial disorder seeking multitherapies. Alzheimer’s Dement. 2010;6(5):420–424. doi:10.1016/j.jalz.2010.04.006

2. Breijyeh Z, Karaman R. Comprehensive Review on Alzheimer ’ s Disease : Published online 2020.

3. Tiwari S, Venkata A, Kaushik A, Adriana Y, Nair M. Alzheimer ’ s Disease Diagnostics And Therapeutics Market. Int J Nanomedicine . 2019;Jul 2019(14):5541–5554.

4. Nelson PT, Alafuzoff I, Bigio EH, et al. Correlation of alzheimer disease neuropathologic changes with cognitive status: A review of the literature. J Neuropathol Exp Neurol. 2012;71(5):362–381. doi:10.1097/NEN.0b013e31825018f7

5. Arnold SE, Hyman BT, Flory J, Damasio AR, Van Hoesen GW. The topographical and neuroanatomical distribution of neurofibrillary tangles and neuritic plaques in the cerebral cortex of patients with alzheimer’s disease. Cereb Cortex. 1991;1(1):103–116. doi:10.1093/cercor/1.1.103

6. Schönheit B, Zarski R, Ohm TG. Spatial and temporal relationships between plaques and tangles in Alzheimer-pathology. Neurobiol Aging. 2004;25(6):697–711. doi:10.1016/j.neurobiolaging.2003.09.009

7. Kentarci K, Knopman DS, Dickson DW, et al. Alzheimer Disease : Postmortem Neuropathologic Correlates of Methods : Results : Conclusion : Radiology. 2008;248(1):210–220.

8. Galimberti D, Scarpini E. Disease-modifying treatments for Alzheimer’s disease. Ther Adv Neurol Disord. 2011;4(4):203–216. doi:10.1177/1756285611404470

9. Porsteinsson AP, Isaacson RS, Knox S, Sabbagh MN, Rubino I. Diagnosis of Early Alzheimer’s Disease: Clinical Practice in 2021. J Prev Alzheimer’s Dis. 2021;8(3):371–386. doi:10.14283/jpad.2021.23

10. Aisen PS, Jimenez-Maggiora GA, Rafii MS, Walter S, Raman R. Early-stage Alzheimer disease: getting trial-ready. Nat Rev Neurol. 2022;18(7):389–399. doi:10.1038/s41582-022-00645-6

11. Guzmán-Vélez E, Diez I, Schoemaker D, et al. Amyloid-β and tau pathologies relate to distinctive brain dysconnectomics in preclinical autosomal-dominant Alzheimer’s disease. Proc Natl Acad Sci U S A. 2022;119(15):1–8. doi:10.1073/pnas.2113641119

12. Long JM, Holtzman DM. Alzheimer Disease: An Update on Pathobiology and Treatment Strategies. Cell. 2019;179(2):312–339. doi:10.1016/j.cell.2019.09.001

13. Fan DY, Jian JM, Huang S, et al. Establishment of combined diagnostic models of Alzheimer’s disease in a Chinese cohort: the Chongqing Ageing & Dementia Study (CADS). Transl Psychiatry. 2022;12(1). doi:10.1038/s41398-022-02016-7

14. Ebenau JL, Timmers T, Wesselman LMP, et al. ATN classification and clinical progression in subjective cognitive decline: The SCIENCe project. Neurology. 2020;95(1):46–58. doi:10.1212/WNL.0000000000009724

15. Meyer PF, Pichet Binette A, Gonneaud J, Breitner JCS, Villeneuve S. Characterization of Alzheimer Disease Biomarker Discrepancies Using Cerebrospinal Fluid Phosphorylated Tau and AV1451 Positron Emission Tomography. JAMA Neurol. 2020;77(4):508–516. doi:10.1001/jamaneurol.2019.4749

16. Hansson O, Lehmann S, Otto M, Zetterberg H, Lewczuk P. Advantages and disadvantages of the use of the CSF Amyloid β (Aβ) 42/40 ratio in the diagnosis of Alzheimer’s Disease. Alzheimer’s Res Ther. 2019;11(1):1–15. doi:10.1186/s13195-019-0485-0

17. Shaw LM, Vanderstichele H, Knapik-Czajka M, et al. Cerebrospinal fluid biomarker signature in alzheimer’s disease neuroimaging initiative subjects. Ann Neurol. 2009;65(4):403–413. doi:10.1002/ana.21610

18. Jellinger A K. Towards a Biological Definition of Alzheimer Disease. Int J Neurol Neurother. 2018;7(1):535–562. doi:10.23937/2378-3001/1410095

19. Grøntvedt GR, Lauridsen C, Berge G, et al. The Amyloid, Tau, and Neurodegeneration (A/T/N) Classification Applied to a Clinical Research Cohort with Long-Term Follow-Up. J Alzheimer’s Dis. 2020;74(3):829–837. doi:10.3233/JAD-191227

20. Jr CRJ, Barkhof F, Bernstein MA, et al. NIH Public Access for Alzheimer ’ s disease. Alzheimers Dement. 2012;7(4):474–485. doi:10.1016/j.jalz.2011.04.007.Steps

21. Chow N, Hwang KS, Hurtz S, et al. Comparing 3T and 1.5T MRI for mapping hippocampal atrophy in the Alzheimer’s disease neuroimaging initiative. Am J Neuroradiol. 2015;36(4):653–660. doi:10.3174/ajnr.A4228

22. Brickman AM, Tosto G, Gutierrez J, et al. An MRI measure of degenerative and cerebrovascular pathology in Alzheimer disease. Neurology. 2018;91(15):E1402–E1412. doi:10.1212/WNL.0000000000006310

23. Schwarz CG. Uses of Human MR and PET Imaging in Research of Neurodegenerative Brain Diseases. Neurotherapeutics. 2021;18(2):661–672. doi:10.1007/s13311-021-01030-9

24. Apostolova LG, Zarow C, Biado K, et al. Relationship between hippocampal atrophy and neuropathology markers: A 7T MRI validation study of the EADC-ADNI harmonized hippocampal segmentation protocol. Alzheimer’s Dement. 2015;11(2):139–150. doi:10.1016/j.jalz.2015.01.001

25. Clarke WT, Mougin O, Driver ID, et al. Multi-site harmonization of 7 tesla MRI neuroimaging protocols. Neuroimage. 2020;206. doi:10.1016/j.neuroimage.2019.116335

26. Wisse LEM, Biessels GJ, Heringa SM, et al. Hippocampal subfield volumes at 7T in early Alzheimer’s disease and normal aging. Neurobiol Aging. 2014;35(9):2039–2045. doi:10.1016/j.neurobiolaging.2014.02.021

27. Wisse LEM, Kuijf HJ, Honingh AM, et al. Automated hippocampal subfield segmentation at 7T MRI. Am J Neuroradiol. 2016;37(6):1050–1057. doi:10.3174/ajnr.A4659

28. Hari I, Adeyemi OF, Gowland P, et al. Memory impairment in amyloid-status Alzheimer’s disease is associated with dentate gyrus atrophy: in vivo MRI at 7T. NeuroImage Clin. 2024;(0):1–27.

29. Jack CR, Bennett DA, Blennow K, et al. NIA-AA Research Framework: Toward a biological definition of Alzheimer’s disease. Alzheimer’s Dement. 2018;14(4):535–562. doi:10.1016/j.jalz.2018.02.018

30. Farlow MR, Andreasen N, Riviere ME, et al. Long-term treatment with active Aβ immunotherapy with CAD106 in mild Alzheimer’s disease. Alzheimer’s Res Ther. 2015;7(1):1–13. doi:10.1186/s13195-015-0108-3

31. Hosseini A et al. Clinical Utility of Cerebrospinal Fluid Aβ42 and Tau Measures in Diagnosing Mild Cognitive Impairment in Early Onset Dementia. J Alzheimer’s Dis. 2022;87(2):771–778.

32. Jak AJ, Bondi MW, Delano-Wood L, et al. Quantification of five neuropsychological approaches to defining mild cognitive impairment. Am J Geriatr Psychiatry. 2009;17(5):368–375. doi:10.1097/JGP.0b013e31819431d5

33. Chertkow H, Feldman HH, Jacova C, Massoud F. Definitions of dementia and predementia states in Alzheimer’s disease and vascular cognitive impairment: Consensus from the Canadian conference on diagnosis of dementia. Alzheimer’s Res Ther. 2013;5(SUPPL. 1):1–8. doi:10.1186/alz190

34. Yushkevich PA, Pluta JB, Wang H, et al. Automated volumetry and regional thickness analysis of hippocampal subfields and medial temporal cortical structures in mild cognitive impairment. Hum Brain Mapp. 2015;36(1):258–287. doi:10.1002/hbm.22627

35. Schuff N, Woerner N, Boreta L, et al. MRI of hippocampal volume loss in early Alzheimers disease in relation to ApoE genotype and biomarkers. Brain. 2009;132(4):1067–1077. doi:10.1093/brain/awp007

36. Vijayakumar A, Vijayakumar A. Comparison of Hippocampal Volume in Dementia Subtypes. ISRN Radiol. 2013;2013:1–5. doi:10.5402/2013/174524

37. Haapalinna F, Paajanen T, Penttinen J, et al. Low Cerebrospinal Fluid Amyloid-Beta Concentration Is Associated with Poorer Delayed Memory Recall in Women. Dement Geriatr Cogn Dis Extra. 2016;6(2):303–312. doi:10.1159/000446425

38. Haldenwanger A, Eling P, Kastrup A, Hildebrandt H. Correlation between cognitive impairment and CSF biomarkers in amnesic MCI, non-amnesic MCI, and Alzheimer’s disease. J Alzheimer’s Dis. 2010;22(3):971–980. doi:10.3233/JAD-2010-101203

39. Stricker NH, Dodge HH, Dowling NM, Han SD, Erosheva EA, Jagust WJ. CSF biomarker associations with change in hippocampal volume and precuneus thickness: Implications for the Alzheimer’s pathological cascade. Brain Imaging Behav. 2012;6(4):599–609. doi:10.1007/s11682-012-9171-6

40. Desikan RS, McEvoy LK, Thompson WK, et al. Amyloid-β associated volume loss occurs only in the presence of phospho-tau. Ann Neurol. 2011;70(4):657–661. doi:10.1002/ana.22509

41. Botha H, Mantyh WG, Graff-Radford J, et al. Tau-negative amnestic dementia masquerading as Alzheimer disease dementia. Neurology. 2018;90(11):e940–e946. doi:10.1212/WNL.0000000000005124

42. Rabinovici GD, Knopman D, Arbizu J, et al. 2024 Updated Appropriate Use Criteria for Amyloid and Tau PET in Alzheimer’s Disease. Butl Hosp Mem Aging Progr. 2024;17:1–71.

43. Tsao A, Sugar J, Lu L, et al. Integrating time from experience in the lateral entorhinal cortex. Nature. 2018;561(7721):57-62. doi:10.1038/s41586-018-0459-6

44. Du AT, Schuff N, Amend D, et al. Magnetic resonance imaging of the entorhinal cortex and hippocampus in mild cognitive impairment and Alzheimer’s disease. J Neurol Neurosurg Psychiatry. 2001;71(4):441–447. doi:10.1136/jnnp.71.4.441

45. Braak H, Braak E. Staging of Alzheimer-Related Cortical Destruction. Int Psychogeriatrics. 1997;9(S1):257–261. doi:10.1017/S1041610297004973

46. Wattmo C, Blennow K, Hansson O. Cerebro-spinal fluid biomarker levels: Phosphorylated tau (T) and total tau (N) as markers for rate of progression in Alzheimer’s disease. BMC Neurol. 2020;20(1):1–12. doi:10.1186/s12883-019-1591-0

47. Hampel H, Blennow K. CSF tau and β-amyloid as biomarkers for mild cognitive impairment. Dialogues Clin Neurosci. 2004;6(4):379–390. doi:10.31887/dcns.2004.6.4/hhampel

48. Vos SB, Winston GP, Goodkin O, et al. Hippocampal profiling: Localized magnetic resonance imaging volumetry and T2 relaxometry for hippocampal sclerosis. Epilepsia. 2020;61(2):297–309. doi:10.1111/epi.16416

49. Düzel E, Berron D, Schütze H, et al. CSF total tau levels are associated with hippocampal novelty irrespective of hippocampal volume. *Alzheimer’s Dement Diagnosis*, Assess Dis Monit. 2018;10:782–790. doi:10.1016/j.dadm.2018.10.003

50. Hampel H, Bürger K, Pruessner JC, et al. Correlation of cerebrospinal fluid levels of tau protein phosphorylated at threonine 231 with rates of hippocampal atrophy in Alzheimer disease. Arch Neurol. 2005;62(5):770–773. doi:10.1001/archneur.62.5.770

51. de Souza LC, Chupin M, Lamari F, et al. CSF tau markers are correlated with hippocampal volume in Alzheimer’s disease. Neurobiol Aging. 2012;33(7):1253–1257. doi:10.1016/j.neurobiolaging.2011.02.022

52. Stav AL, Johansen KK, Auning E, et al. Hippocampal subfield atrophy in relation to cerebrospinal fluid biomarkers and cognition in early Parkinson’s disease: a cross-sectional study. npj Park Dis. 2016;2(1). doi:10.1038/npjparkd.2015.30

53. Devi G, Scheltens P. Heterogeneity of Alzheimer’s disease: Consequence for drug trials? Alzheimer’s Res Ther. 2018;10(1):1–3. doi:10.1186/s13195-018-0455-y

54. Jack CR, Hampel HJ, Universities S, Cu M, Petersen RC. A new classification system for AD , independent of cognition A / T / N : An unbiased descriptive classification scheme for Alzheimer disease biomarkers. Neurology. 2016;0(July):1–10.

